# Outcomes following short hospital stay after total knee replacement in a regional setting: A prospective analysis of an observational cohort in a public hospital treated 2018 - 2019

**DOI:** 10.1101/2020.03.08.20031989

**Authors:** Manaal Fatima, Corey Scholes, Amanda Tutty, Milad Ebrahimi, Michel Genon, Samuel J. Martin

**Affiliations:** EBM Analytics, Crows Nest, NSW, Australia; Grafton Base Hospital, Northern NSW Local Health District, Grafton, NSW, Australia

**Keywords:** length of stay, enhanced recovery, knee arthroplasty, regional hospital

## Abstract

Functional outcomes and patient satisfaction following short length of stay (LoS) after total knee arthroplasty (TKA) in the regional context have not been explored. This study aimed to report on complications, functional outcomes and satisfaction of patients discharged from acute (≤2 days) stay, up to 6 weeks after TKA in a regional hospital.

Patients were prospectively recruited from August 2018 to August 2019. Demographic data, intraoperative factors and the incidence of complications and emergency department (ED) presentations were retrieved from hospital records. Preoperative and 6 weeks postoperative assessments collected range of motion (ROM), timed up and go (TUG), 6 minute walk test (6MWT) and total Oxford knee score (OKS), with patient satisfaction assessed at follow-up via a questionnaire. A directed acyclic graph approach was used to perform logistic and linear regression to assess relationships between patient and treatment characteristics with short-term outcomes.

Median LoS for the period was 2 days, influenced by age, gender, bilateral status and body mass index. A shorter LoS was significantly associated with functional outcomes and patient satisfaction, with 74.4% of patients satisfied with their knee and 88.4% satisfied with their LoS. At six weeks, significant improvements in all scores were found, however only the change in OKS exceeded the minimal clinically important difference (MCID) threshold. Patient satisfaction correlated with change in TUG exceeding MCID. Complications at 6 weeks post surgery were affected by Aboriginal and Torres Strait Islander status, marriage status, smoking history, history of chronic pain and mental health conditions, and the number of ED presentations was associated with preoperative TUG and comorbidities.

The findings establish that short LoS after TKA in a regional hospital is associated with good outcomes and a high patient satisfaction, but patient factors and comorbidities associated with an increased risk of complications and ED presentations should be considered for postoperative management and patient care.

## Introduction

Total joint arthroplasty is a high-cost surgery, with direct expenditure exceeding $AUD1 billion annually in Australia ^1^ and projected to reach $AUD5.3 billion per annum in the next decade ^2^. Sociodemographic factors, perioperative characteristics and postoperative complications, particularly surgical site infections, increase costs of total knee arthroplasty (TKA) by up to 60%

^1^. There are a number of factors affecting LoS that need to be considered in the context of postoperative hospital stay. These include age, sex, marital status, American Society of Anesthesiologists (ASA) score, comorbidities and complications ^3^, as well as wound exudate and range of motion (ROM) at day 0 ^4^. A higher preoperative body mass index (BMI) and poor VR-12 Physical Component Score reportedly increase LoS, as do patient factors such as ethnicity, lower household income and bilateral status ^5,6^.

Modifiable strategies to reduce costs associated with medical services such as TKA include the implementation of enhanced recovery protocols (ERPs) to expedite hospital discharge. In Australia, the average hospital length of stay (LoS) is 5.2 days in private and 5.7 days in public hospitals ^7^. ERPs aim to maximise efficiency by reducing LoS to ≤2 days whilst maintaining an acceptable level of patient outcomes and satisfaction, and have gained traction with their successful implementation ^8^. Patient satisfaction and outcomes in ERP programs are typically favourable ^9^, with patients reporting lower pain scores at rest and with activity despite significantly lower morphine utilisation, with no difference in readmissions at 30 days ^10^. Data on patient experience and satisfaction however, are limited, and while it appears that patients are generally satisfied with their ERP experience, their real concerns may be masked by acceptance of early discharge and eagerness to return home to family, rather than be confined to a hospital bed ^11^.

Limited studies have reported on patient reported outcome measures (PROMs), but have found that quality of life was not compromised after hip or knee elective surgery, and that scores continued to improve up to 12 months following surgery with an ERP ^11^. Most postoperative follow-ups in studies reporting on functional outcomes have typically been conducted at 3-4 months ^8,11^, though complications and readmissions generally occur within 30 days of surgery ^1^. A systematic review and meta-analysis reported reduced incidence of postoperative complications and 30-day mortality rate in ERP groups, but no difference in groups for 30-day readmissions. With short-term follow-up after TKA, studies have reported a decrease in pain and improved function in the first 7 days ^12^, up to 3 weeks ^13^ and at 6 weeks ^9^ postoperatively in patients discharged from an ERP program. ERPs in the regional setting are not as frequently explored, but LoS can be successfully reduced over time without significant increases in the rate or severity of complications or representation to acute care and subsequent readmission ^14,15^. In an Australian regional hospital, a retrospective review revealed that LoS was significantly reduced from 6 to 2 days over a period of 6 years ^14^, and in an American rural setting, LoS was reduced from 3.7 to 2.8 days over a 4 year period ^15^. While representation to emergency and hospital readmission remained low for the regional hospital ^14^, it remains unclear whether the reductions in LoS can be maintained over time, or whether satisfactory patient outcomes and experience can be achieved with a reduced stay in a regional population.

To address these concerns, a prospective analysis of patients receiving TKA with a reduced hospital stay in a regional setting is required. Therefore, the aims of this study were to: 1) report on the incidence of complications up to 6 weeks follow-up in patients undergoing TKA at a regional hospital where LoS had been reduced ^14^, and 2) report on the functional outcomes and satisfaction of this cohort discharged from acute stay through an ERP.

## Methods

### Patient selection and informed consent

A prospective analysis of an observational cohort with census sampling and exclusion criteria was performed on all patients receiving a total knee replacement (TKR) under the care of one of two surgeons (SM or MG) between August 2018 to August 2019. Ethical approval was obtained from the North Coast Human Research Ethics Committee (2018/ETH000120), and informed consent was obtained prior to surgery. Patients were included if they consented for a TKA performed at Grafton Base Hospital, were older than 18 years, discharged at the time of data collection, and did not meet any of the exclusion criteria. Exclusion criteria included patients requiring a multistage revision, surgeries not performed by the authors and patients who received a preoperative diagnosis other than degenerative arthritis. Patients with a clinical diagnosis of a mental or neurological illness that precluded retrieval of feedback via interview, paper or electronic questionnaires were excluded from PROMs retrieval but were included for the analysis of other outcomes. The number of patients eligible for the study after the application of the inclusion and exclusion criteria is shown in **Figure 1**.

**Figure 1:**
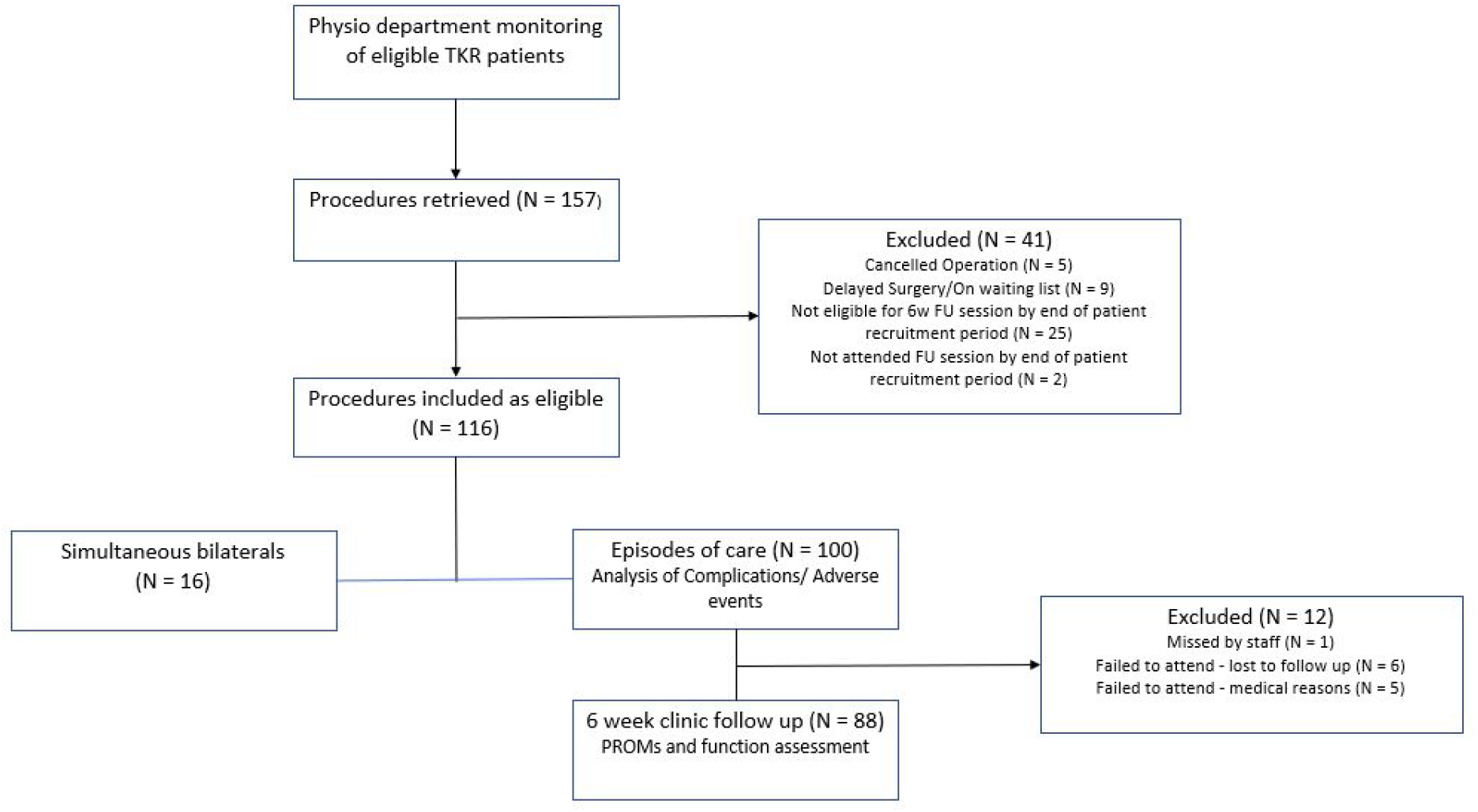
STROBE ^16^ diagram illustrating patient inclusion and analysis for the study.

### Surgical technique

Spinal anesthetic with local intra-articular infiltration was used for the majority of cases, with some patients requiring additional general anesthetic as determined by the anesthesiologist. Patients received a single dose of prophylactic antibiotics just prior to induction. A subvastus approach was used by both participating surgeons during the study period. Tourniquet use was used sparingly, with tranexamic acid administered intravenously in anaesthetic bay prior to surgery, as well as topically during the procedure. Tranexamic acid was continued orally or intravenously in the immediate postoperative period. Patients received a fixed bearing TKA (Stryker, Kalamazoo, MI), under computer navigation guidance (ASM, Stryker, Kalamazoo, MI) with one surgeon routinely performing a cruciate retaining knee replacement and rarely resurfacing the patella, and the other a posterior stabilized replacement with routine patella resurfacing. The majority of patients received anaesthetic delivered by intraarticular injection consisting of 0.2% naropin at a dose determined by the anaesthetist and addition of either betamethasone (5.7mg) or triamcinolone (40mg) depending on availability. Closure was undertaken in layers including the synovial layer. Later in the series a small number of cases received continuous intraarticular infusion with 270mL ropivacaine 0.2% at a speed of 5 mL/hour through an elastomeric infusion pump (Painbuster, I-Flow Corp, Lake Forest, CA). The pump was removed on day 2 by the nursing staff post discharge. Bandaging with Coban was performed directly over the sterile dressing with no padding. All patients received prophylaxis against deep vein thrombosis. All patients received standardized physiotherapy in accordance with the institution’s knee arthroplasty discharge pathway, and patients were considered for discharge once mobile, medically stable, and with confirmed discharge destination as per standard. Hospital discharge date was patient, nurse and physiotherapist initiated for patients who had been medically cleared.

### Data collection

Patients were prospectively recruited for inclusion into the study. A dataset was prepared prior to data collection outlining the required variables and the location of the planned data fields with input from clinical investigators, the head of department of hospital medical records, and allied health staff. A chart review was performed as previously described ^14^, but with a new team to retrieve intraoperative factors such as analgesia and anesthesia strategy, as well as complications, emergency department (ED) presentations and hospital LoS. Patient factors retrieved from electronic medical records included age, gender, body mass index (BMI), financial status,marital status and whether or not the patient identified as Aboriginal and/or Torres Strait Islander. Mental health status was collected with binary responses (yes or no), identifying patients with a diagnosed (yes) mental health condition such as depression, anxiety or schizophrenia. Patient comorbidities were extracted as free text and re-coded into binary indicator variables (**Supplementary table A1**). All patients underwent a physiotherapy assessment preoperatively and at 6 weeks follow-up clinic to record data on range of motion (ROM) in flexion and extension, timed up and go (TUG), 6 minute walk test (6MWT) and total Oxford knee score (OKS) using a paper form. A questionnaire was additionally administered on paper at postoperative follow-up to assess patient satisfaction with their knee replacement and their hospital LoS, with patients providing a numerical rating out of 10 (**Supplementary material B**). Questionnaires were scored by hand and transferred to a spreadsheet for further analysis.

### Statistical analysis

Data extracted from chart review was recorded in preparation for statistical analysis (**Supplementary table A1**). Descriptives were calculated for each variable and visualised with probability distribution plots (continuous variables) and cross-tabulation (categorical variables) to identify outliers or transcription errors. Length of stay was calculated as the difference in days between the date of admission to the date of discharge and was visualised over time with a line plot to assess stability over the recruitment period. Complications were graded for severity using the modified system described by Sink et al ^17^. A positive change of 3.4seconds or above was used to define the minimal clinically important difference (MCID) for the timed up and go test (TUG) ^18^. A positive change of 55.4 metres or above was used to define MCID for the 6-minute walk test (6MWT) ^19^. A positive change of 5 points or above was used to define MCID for the OKS ^20^. Stepwise (forwards-backwards) linear regression was used to establish relationships between patient characteristics, comorbidities, treatment characteristics and outcomes (length of stay, complications, ED presentations, MCID of function scores; **Supplementary table A2**). Length of stay was transformed to satisfy the assumption of normality in model residuals using a Box-Cox transformation with optimal lambda. An alpha of 0.1 was used to include/exclude variables from the model and model fit was assessed with adjusted R^2^. Coefficients with standard error were reported for each factor identified in the final model (continuous variables) or odds-ratios with 95% confidence intervals (categorical variables). The relationship between length of stay and satisfaction scores were assessed with univariate correlation analysis (Spearman rho). All statistical analyses were performed in dedicated software (Minitab v18, Minitab Inc, USA). A directed acyclic graph approach ^21^ was used to summarise relationships identified in the statistical analysis using an open-access R package ^22^.

## Results

### Patient characteristics

The patient sample of 100 presentations operated from 1 August 2018 to 12 August 2019 were aged 68 (median, IQR 62 - 75.8) and 54% female, with a BMI of 32.9 (IQR 27.8 - 38.5) and 16% underwent simultaneous bilateral procedures. Patients presented with a median of 2 comorbidities (IQR 1 - 3) with the majority presenting with a cardiovascular condition (72%), followed by a history of smoking (20%), previous arthroplasty surgery on another joint and other musculoskeletal conditions (17%), respiratory conditions (16%), diabetes (14%), spinal conditions (12%), chronic pain (11%), history of cancer (11%) and neurological conditions (7%). Patients being treated for a mental condition (e.g. depression, anxiety) made up 12% of the cohort and *other* conditions not listed affecting 10%. Six patients (6%) identified as being of Aboriginal and/or Torres Strait Islander descent.

### Length of Stay

Median LoS was 2 days (IQR 2 - 4), with 58% discharged by day 2 and 16% staying longer than 5 days. LoS was not completely stable throughout the monitored period (**Figure 2**), with a trend (P = 0.01) depicted by a *bathtub* curve (increased, stable, increasing). A linear model identified older patients, female gender, diabetes comorbidity and bilateral procedures as being associated with significantly longer length of stay (**Table 1**). A previous history of arthroplasty in other joints was significantly associated with reduced length of stay (**Table 1**).

**Table 1:** Summary of factors associated with length of stay (transformed)

| | $\beta$ | Standard Error | P - value |
| --- | --- | --- | --- |
| **BMI** | 0.03 | 0.02 | 0.09 |
| **Age at surgery** | 0.05 | 0.02 | 0.001 |
| **Female** | 0.12 | 0.07 | 0.001 |
| **Bilateral** | 0.11 | 0.03 | 0.02 |
| **OKS preoperative** | -0.04 | 0.02 | 0.02 |

**Figure 2:**
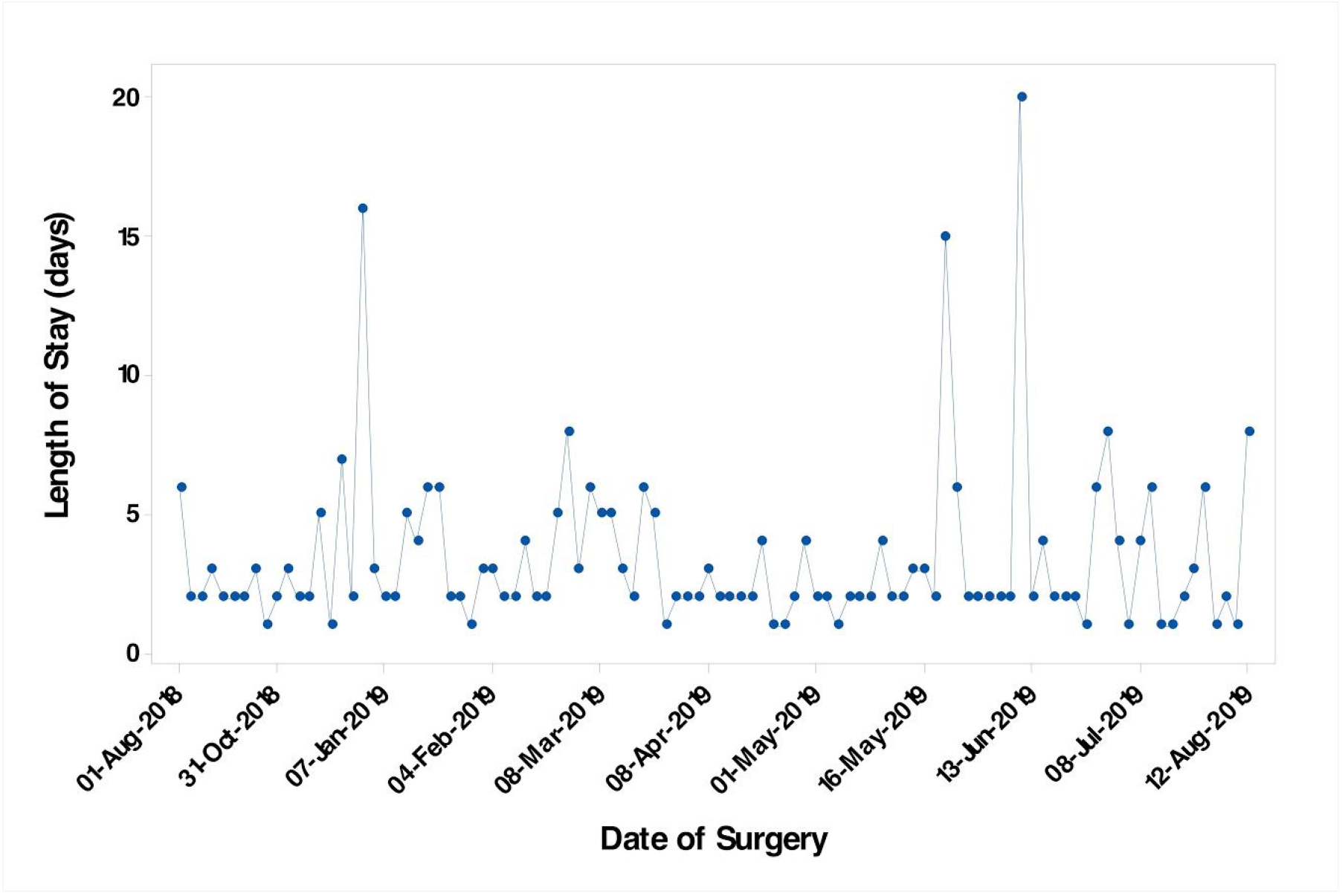
Potential trend in length of stay (bathtub curve)

### Complications, ED presentations and readmissions

A total of 3 patients had Grade 1 complications noted during in-hospital stay. A proportion of patients (N = 27) presented to the ED (total = 37) (median presentations = 1; IQR 1 - 3, maximum = 4) at a median of 7 days (IQR 2 - 13) between discharge and first presentation (Grade I = 4; Grade II = 16; Grade III = 10). Of those, 10 patients were readmitted for revision due to fracture (1), washout (1), wound closure (1) and upper limb fracture fixation (1), while the rest were discharged after observation, discharged with home monitoring or treated with precautionary antibiotics. The presence of a complication up to 6 weeks after discharge was significantly associated with a longer distance for preoperative 6MWT, identifying as Aboriginal and/or Torres Strait Islander, a history of smoking, or a notable mental condition (**Table 2**). Bilateral procedures were at significantly reduced risk of presenting with a complication (**Table 2**). The number of ED presentations was associated with preoperative TUG, a history of diabetes and a history of smoking (**Table 3**).

**Table 2:** Summary of factors associated with a *graded* complication

|  | **Odds Ratio** | **Confidence Interval** | **P- Value** |
| --- | --- | --- | --- |
| Aboriginal and/or Torres Strait Islander peoples | 13.8 | 2.3 - 83.9 | 0.001 |
| Bilateral procedure | 0.07 | 0.01-0.35 | <0.001 |
| Smoking history | 7.1 | 2.8 - 17.9 | <0.001 |
| Mental health conditions | 5 | 1.7 - 14.4 | 0.009 |
| Preoperative 6MWT distance | 1.004 | 1.0 - 1.007 | 0.027 |

**Table 3:** Summary of factors associated with number of emergency department presentations

| | $\beta$ | Standard Error | P - value |
| --- | --- | --- | --- |
| Preoperative TUG | -0.05 | 0.22 | 0.022 |
| Aboriginal and/or Torres Strait Islander peoples | 1.07 | 0.49 | 0.052 |
| Diabetes | 0.73 | 0.44 | 0.114 |
| Smoking history | 1.08 | 0.38 | 0.005 |
| Bilateral status | -1.18 | 0.74 | 0.057 |

### Functional outcomes and satisfaction

On average, ROM changed little between preoperative assessment and the 6-week follow up, with significant improvements in OKS, 6MWT and TUG (**Table 4**). However, only the change in OKS exceeded the MCID threshold. An individual, anchor-based responder analysis revealed 73.5% of the sample reported a change in OKS that exceeded the MCID threshold, with just 4.9% reporting a worse score >MCID. In addition, 36.4% improved >MCID for 6MWT, while 14.8% worsened. For the TUG, 21.6% improved and 6.8% worsened. The presence of certain comorbidities, marriage status and presence of a complication were significantly associated with the probability of a positive MCID change in these functional measures (**Table 5**). Of the patients that responded to the satisfaction questionnaires (N = 86), 74.4% of patients were satisfied (8/10 or above) with their knee (median 9/10, IQR 7 - 10) and 88.4% were satisfied with their length of stay (median 10, IQR 9 - 10). A shorter LoS was not associated with the satisfaction rating of the knee (out of 10) (rho = −0.05, P = 0.68), but was moderately inversely associated with the satisfaction rating of LoS (rho = −0.22, P = <0.001). A summary of relationships illustrated that while patient length of stay was influenced by patient factors (age,gender, preoperative function, comorbidities) it was not directly associated with any functional outcomes other than satisfaction (**Figure 3**).

**Table 4:** Summary of functional outcomes at preoperative and postoperative timepoints

|  | Preoperative |  |  | Postoperative |  |  |  |
| --- | --- | --- | --- | --- | --- | --- | --- |
|  | N | Median | IQR | N | Median | IQR | One-sample t-test P-value |
| Fixed flexion | 116 | 4 | 0-10 | 102 | 1.5 | 0-5 |  |
| Max Flexion | 116 | 115 | 100-125 | 102 | 105 | 100-115 |  |
| OKS | 116 | 18.5 | 14.3 - 25 | 102 | 31.5 | 26.8 - 37 |  |
| OKS delta | - | - | - | 102 | 11<sup>a</sup> | 3.8 - 19 | <0.001 |
| 6MWT (m) | 100 | 325 | 234-400 | 88 | 338 | 275-425 |  |
| 6MWT delta |  |  |  | 88 | 3<sup>b</sup> | -25 - 90 | 0.002 |
| TUG (s) | 100 | 10.6 | 8.3 - 13.6 | 88 | 11.6 | 9.8 - 21 |  |
| TUG delta |  |  |  | 88 | 0.24<sup>c</sup> | -1.5 - 3 | 0.05 |

<sup>a</sup>Above MCID of 5 points; <sup>b</sup>Below MCID of 54 metres; <sup>c</sup>below MCID of 3.4sec

**Table 5:** Summary of anchor-based responder analysis

|  | Outcome (%) | Model R<sup>2</sup> (%) | Predictors | OR (95%CI) | P-value |
| --- | --- | --- | --- | --- | --- |
| **OKS MCID\*** | Worse (26.5) | 5.2 | No previous arthroplasty | 3.4 (1.2 - 9.6) | 0.02 |
|  |  |  | CM - Pain | 4.1 (1.01 - 16.9) | 0.032 |
|  |  |  | CM - Respir | 2.4 (0.9 - 6.1) | 0.06 |
| **6MWT MCID** | Improved (36.4) | 2 | CM - Spinal | 3.2 (1.05 - 8.4) | 0.027 |
| **TUG MCID** | Improved (21.6) | 18.3 | Female | 6.3 (2.7 - 14.5) | <0.001 |
|  |  |  | Married | 3.1 (1.4 - 7.0) | 0.005 |
|  |  |  | CM - Diabetes | 6.6 (1.5 - 18.6) | <0.001 |
|  |  |  | Unilateral surgery | 13.5 (3.2 - 56.3) | <0.001 |

\* Model outcome set as <MCID; CM - comorbidity

**Figure 3:**
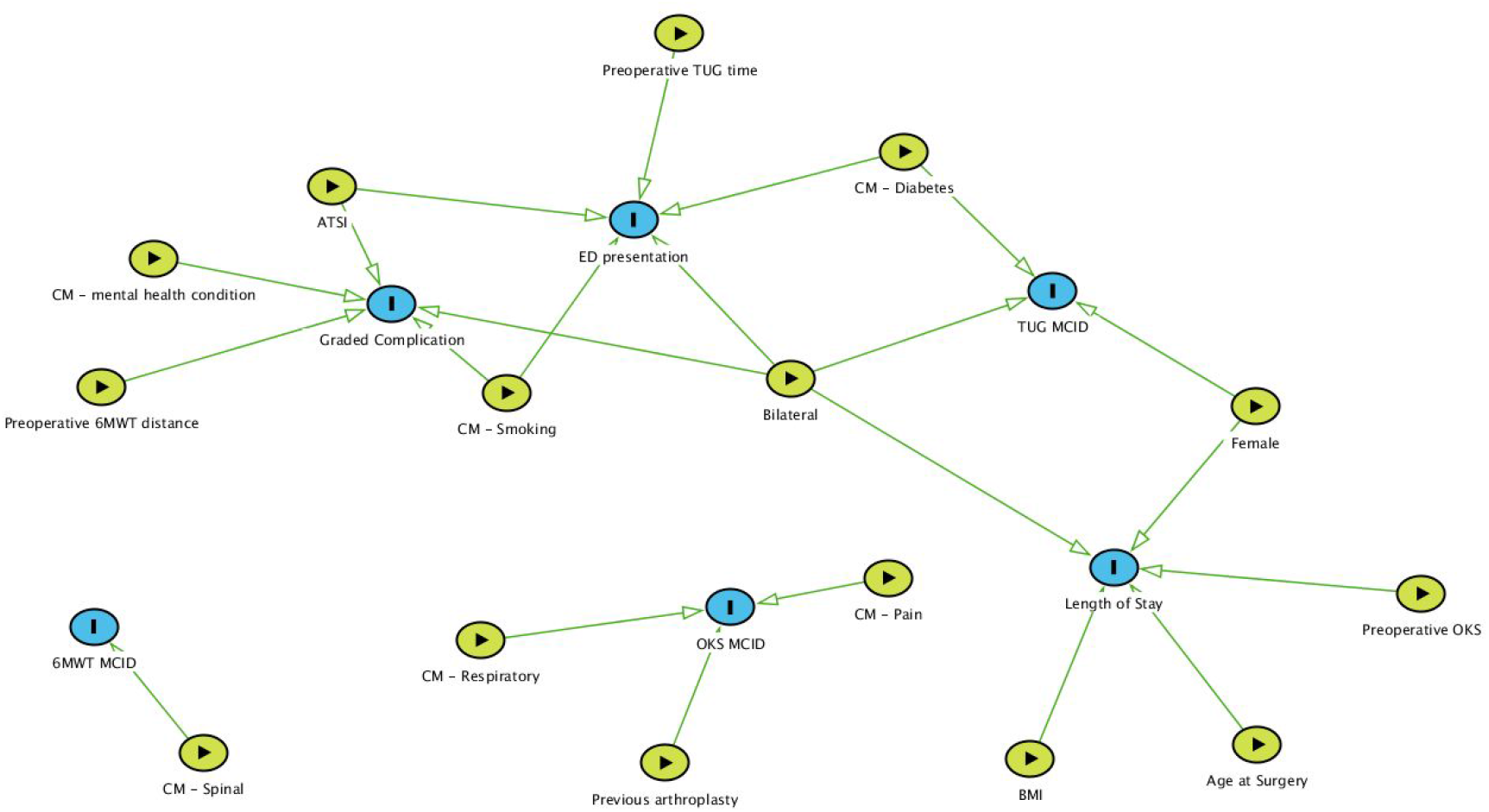
Summary of significant relationships (green directional arrows) between exposures (green circles) and outcomes (blue circles) illustrated with a directed acyclic graph approach. CM - comorbidity; ATSI - Aboriginal or Torres Strait Island peoples; MCID - minimal clinically important difference; ED - emergency department

## Discussion

In the regional context, the short-term incidence of complications and adverse events following short hospital LoS after TKA, as well as functional outcomes and patient satisfaction have not been explored. This study aimed to investigate the incidence of complications up to six weeks follow-up in patients undergoing TKA at a regional hospital, and identify the factors associated with functional outcomes and patient satisfaction. The findings of this study establish that a short LoS after TKA in a regional hospital can be achieved with good functional outcomes and a high level of patient satisfaction, but certain patient factors and comorbidities need to be considered to minimise the risk of complications and ED presentations up to six weeks post-surgery.

While the median LoS in this study was 2 days, as previously reported ^14^, LoS was not maintained uniformly over the course of a year. The reasons for loss of stability in the average LoS over the study period remain to be identified. The present study identified older age, female gender ^3,14^, bilateral surgery, higher BMI ^5^ and lower preoperative OKS associated with increased hospital LoS in agreement with contemporary literature. However, these findings may reflect bias in the discharge process within the department in contrast to actual risk factors associated with patient safety post-discharge or a tendency for these patients to elect to stay longer.

The short LoS (≤2 days) was positively associated with functional outcomes and patient satisfaction, with significant improvements in all scores (OKS, 6MWT and TUG) found, however only the change in average OKS exceeded the MCID threshold. The OKS reported in this study was lower preoperatively (18.5 vs 23) and at 6 weeks postoperatively (31.5 vs 35) ^9^, but comparable to the postoperative change in TUG (0.24 v −1.7) at the same length of follow up ^12^. A lower preoperative OKS was also associated with longer LoS in this study, which highlights potential for preoperative OKS as a predictor of extended LoS, but warrants further work.

An important finding in this study was that the incidence of hospital readmissions (10% vs. 3%) and ED presentations (27% vs 6%) were much higher, even though the incidence of graded complications was lower (20% vs. 25%) compared to our previous retrospective analysis ^14^. Anecdotally, patients reported a 3-4 week wait for access to primary care, which is likely to account for the high presentation to the ED. In New South Wales (NSW), Australia, 33-39% have difficulty accessing out-of-hours medical care, compared to 17% in major cities ^23^. In regional NSW, patients had a 70% chance of getting an appointment with a general practitioner (GP) within two weeks, and a 30% chance of not getting anything ^24^. While the probability of seeing a GP on the same day was highly variable (11-62%), it was still lower than metropolitan areas (78%).

Targeted home-based hospital care that addresses risk areas such as wound management may provide a cost-effective option to reduce unplanned ED presentations and readmissions ^25,26^. Improving access to primary care by encouraging patients to book a GP appointment for one week after their surgery and improving preoperative education as well as postoperative clinical management are strategies that may supplement ERP programs to reduce ED presentations and readmissions in the weeks after surgery.

While ERPs present a strategy to reduce costs associated with major elective surgery, a high incidence of ED presentations may strain hospital resources. The present results can be used to prioritise patients for intervention to reduce adverse events and allocate resources to reduce the incidence of ED presentations and readmissions in the first one to two weeks after hospital discharge. Patients identifying as Aboriginal or Torres Strait Islander, unmarried, with a history of smoking, diabetes or a mental health condition and longer preoperative 6 minute walk test result or shorter preoperative TUG, were at increased risk of adverse events in the present series. Aboriginal and/or Torres Strait Islander peoples represented 6% of the study cohort, close to the proportion identified for Grafton (8.7%) in the 2016 census, which is 3 times higher than the proportion at the state and national levels ^27^. This is an important consideration that needs to be taken into account for ERP programs in regional Australia, particularly where the proportion of Aboriginal and/or Torres Strait Islander peoples is higher. An increased risk of complications and readmissions has previously been attributed to patient comorbidities ^28^. Hospital LoS was not associated with adverse events in this study in agreement with previous studies ^10,28^. In addition, the majority of ED presentations were for minor issues that in all likelihood could have been dealt with by an appropriate primary care provider, and the use of a screening tool that includes physical function assessment with TUG or 6MWT could be considered in conjunction with a home-based care model.

A high level of average satisfaction was reported this cohort, with 74.4% of patients satisfied with their knee and 88.4% satisfied with their LoS. High satisfaction ratings are commonly reported for patients in an ERP program ^10,11^, but this investigation is the first to demonstrate a moderate association between patient satisfaction with their LoS, and their length of stay. The observation of patient satisfaction in the presence of functional improvement suggests that patients are not merely satisfied with early discharge because they are able to return home ^11^, but further verification is required to establish whether their feeling of satisfaction is associated with a quantifiable improvement in their knee function.

This study is the first to explore complications, functional outcomes and patient satisfaction following TKA with an ERP program in a regional hospital, however, the findings must be interpreted in light of certain limitations. While the postoperative follow-up in the study is short at 6 weeks, complications and readmissions generally occur within 30 days of surgery ^8,11^ and functional improvements can be captured within 7 days, with no significant differences between fast-track and regular discharge groups observed at 1-5 years ^12^. However, the MCID cut-off for the functional tests (TUG, 6MWT and OKS) is at 6 months rather than 6 weeks ^19^, and changes at short-term follow-up may be too volatile and variable between cases. Higher ED presentations in the short-term due to restricted access to primary care has important implications for the regional setting. For the Australian context, Aboriginal and/or Torres Strait Islander peoples were significantly at risk of complications at 6 weeks, but there may be limited ability to generalise this across the country, as the proportion of Aboriginal and/or Torres Strait Islander peoples residing in Grafton is 3 times higher than state and national levels. More broadly, the ability to generalise the results across hospitals with more surgeons and in rural settings with more restricted services and facilities remains unknown.

## Conclusion

Short LoS after TKA in a regional hospital can be achieved without compromising patient-reported and functional outcomes, whilst maintaining a high level of patient satisfaction with both the surgery and the subsequent LoS. The presence of comorbidities, having a lower preoperative TUG, and being of Aboriginal and/or Torres Strait Islander descent are key factors associated with a risk of complications and ED presentations, and should be considered in the context of postoperative management and patient care in a regional ERP program.

## Data Availability

Blinded data may be requested from the corresponding author subject to reasonable use.

## Acknowledgements

The authors would like to acknowledge Joanne Martin, Dominic Bullock, Allison Lollback, Mac Cowley, and Meredith Harrison-Brown for their assistance with data retrieval and manuscript preparation.

